# Evaluating Cardiometabolic Indices for Predicting Thrombotic Risk in the Indian Adult Population: A Comparative Analysis

**DOI:** 10.1101/2025.07.18.25331803

**Authors:** Hariharan Seshadri, Yogesh Subramanian, Naveenkumar Nallathambi, R. Krithikavalli, C. Hariharan

**Author notes:** **Corresponding author**, ***Hariharan Seshadri,* M.B.B.S.,** Institute of Internal Medicine, Madras Medical College and Rajiv Gandhi Government General Hospital Chennai, Tamil Nadu, India, Email ID.

## Abstract

**Background:** Cardiometabolic indices are quick measures of visceral adipose dysfunction that bear significant association with risk of future thrombotic events. Traditionally, body mass index (BMI) and waist circumference (WC) have been used for screening large population masses. With the advent of novel indices with better predictive value, it is imperative to validate the utility of these indices in a demography-specific fashion.

**Objective:** In our study, we have attempted to identify the best-performing predictor and its optimal cut-off point specific to the Indian population.

**Methods:** Our study is a hospital-based cross-sectional study involving 200 participants visiting the master health check-up center of our tertiary-care institution. After clinical, anthropometric and biochemical assessments, twenty cardiometabolic indices were calculated for all the participants. Individual cardiovascular, stroke and thromboembolic risks were ascertained through Framingham Risk Score, CHA_2_DS_2_VASc score, and Padua Prediction Score, respectively. ROC analysis was done to identify the indices with statistically significant predictive association for thrombotic risk.

**Results:** In our study, eight indices (NVAI, METS-VF, ABSI, CI, WHR, CVAI, TyG-WC, CMI) have shown significant predictive potential for coronary risk, and eleven indices (RFM, BFP, NVAI, WHtR, BRI, TyG-index, CI, VAI, ABSI, TyG-WC, METS-IR) for stroke risk, while four indices (NVAI, ABSI, CI, TyG-WC) are predictive of both risks.

**Conclusion:** Overall, NVAI is the best predictor of future thrombovascular catastrophe for the Indian demography (FRS AUC = 0.769, CHA_2_DS_2_VASc AUC = 0.663; p-value < 0.05) with optimal cut-off point of 0.972 (males = 0.993, females = 0.913). BMI has shown poor predictive performance for thrombotic risk as compared to the other indices. Cardiometabolic indices, in general, have limited utility in prediction of future venous thromboembolic events.

## 1. INTRODUCTION

Coronary artery disease, stroke and venous thromboembolism are devastating, yet common causes of mortality and morbidity across the globe in the modern era. The rising trends of diabetes mellitus, hypertension, dyslipidemia and obesity, alongside poor dietary choices and sedentary lifestyle have set the stage for the rampant escalation in the incidence of such thrombotic catastrophes in the recent times (1).

At the very core of these entities lies adipose tissue dysfunction (the basis for metabolic syndrome), stemming from the complex biochemical orchestration of genetic and lifestyle factors(2). The subsequent aberrations in adipokine levels induces endothelial dysfunction, resulting in a pro-inflammatory (with elevated interleukin - 6, 8, 18, and tumor necrosis factor-alpha levels) and a pro-thrombotic state characterized by enhanced platelet activation, elevated levels of von Willebrand factor, Factor VII and VIII, fibrinogen, and plasminogen activator inhibitor (PAI)-1, thus leaving the affected individuals susceptible to thrombotic sequelae in the future (3).

Decades of scientific research have been invested to identify modalities for ascertaining the ‘lipotoxicity’ associated with metabolic syndrome with the hope of obtaining predictive insights into the future risk of such thrombotic complications. Several studies have pointed at adipose tissue mass and distribution (subcutaneous and visceral adipose tissue) as useful predictors, with visceral adiposity having a greater correlation with the metabolic derangements (4). Although only CT, MRI and Dual-energy X-ray absorptiometry can provide a direct volumetric estimate of the adiposity and its distribution, alternative measures based on anthropometry and biochemical profile have gained prominence due to the limited utility of the imaging modalities in screening the large masses (5).

The body mass index (BMI) has been the most commonly used surrogate marker for quantifying adiposity in diverse populations. However, the short-comings of BMI in differentiating between lean mass and fat mass, and describing body fat distribution have paved the way for work on novel indices to mitigate these issues. Waist circumference (WC), Waist-hip ratio (WHR) and Waist-height ratio (WHtR) have been shown to have a stronger correlation with visceral adiposity than BMI (6). The Cardiometabolic index (CMI), Visceral adiposity index (VAI), Chinese visceral adiposity index (CVAI), and New visceral adiposity index (NVAI) are markers of adipose function and metabolic health with predictive potential for diabetes and 10-year atherosclerotic cardiovascular disease (ASCVD) risk (7–10). A body shape index (ABSI) and Conicity index (CI) have displayed good discriminatory potential for abdominal adiposity and all-cause mortality in the previous studies (11,12). Triglyceride-glucose (TyG) index, TyG-WC, TyG-BMI, and Lipid accumulation product (LAP) are reliable surrogates for insulin resistance, visceral adiposity, diabetes and ASCVD risk (13,14).

Population-specific validation studies in different demographic regions [*Liu et al.* (15)*, Huang et al.* (16)] have revealed that the predictive potential of these indices is variable in different population groups, and the choice of the right indices for clinical application must be made on the basis of validation studies on the residing population. However, there is a dearth of such comparative evidence specific to the Indian population, where the prevalence of metabolic syndrome is between to 28% to 33% (in contrast to the 12.5 – 31.4% prevalence worldwide, as of 2022) (17,18). Our study seeks to bridge this major lacuna in the medical literature. In particular, we seek to re-evaluate the utility of BMI in routine clinical practice, and seek to suggest a better surrogate marker for effective large-scale screening of thrombotic risk.

Alongside the traditional cardiovascular and cerebrovascular complications that have been widely evaluated in the previous studies, we also seek to assess the utility of cardiometabolic indices in predicting the risk of venous thromboembolism as a theoretical consequence of the pro-thrombotic state furnished by the adipocyte dysfunction (19).

## 2. SUBJECTS, MATERIALS AND METHODS

### 2.1. STUDY DESIGN AND PARTICIPANTS

This study is a hospital-based cross-sectional study conducted between April to June 2023 at Rajiv Gandhi Government General Hospital, Chennai (a tertiary care center in India). The adult population visiting the hospital’s Master Health Check-up Center (a Government-subsidized general health screening initiative for the common public) during the study period was thoroughly scrutinized and individuals satisfying the selection criteria were approached and recruited into the study.

We included adult participants in the age group of 30 – 74 years by consecutive sampling. Participants <30 or >74 years of age were not included as the Framingham Risk Score for cardiovascular disease is not applicable for this age group. Our exclusion criteria comprised of all patients who did not consent to participate, and all medical conditions that cause severe distortion of the anthropometry and/or biochemical parameters of the participants in proportions that are unexplainable by obesity alone, such as pregnancy, ascites, severe hepatic/renal disorders, injuries, and congenital anomalies.

The sample size of the study population was 200. This was obtained by rounding-off the required sample size of 192, calculated using the area under ROC curve for TyG-WC index of 0.683 (16) with a prevalence of coronary artery disease of 12% (20) at α-error = 0.05 and β-error = 0.2.

The study was commenced after obtaining approval from the Institutional Review Board and Ethics Committee of Madras Medical College and Rajiv Gandhi Government General Hospital, Chennai (Reference no.: 28032023) and executed in accordance to the Declaration of Helsinki. Participants were included in the study only after gaining their consent for participation. Written informed consent was obtained from all study participants after explaining the nature, purpose and procedure of the study.

### 2.2. DATA COLLECTION

A detailed clinical history was elicited from the participants through which demographic details, history of comorbid conditions, medications usage, and smoking habits was obtained. All clinical interviews were conducted by the principal investigator to avoid inter-observer variance, and interview time was maintained the same among different patients.

Height (cm) of the participants was quantified using a wall-mounted scale after ensuring an erect posture with shoes removed. Waist circumference (cm) was measured using a non-elastic tape with participants wearing clothing in standing position, along the horizontal plane midway between costal margin and iliac crest after normal expiration (WHO/IDF recommendation). Hip circumference (cm) was measured similarly along the widest portion of the gluteal region in a plane parallel to the floor. All length measurements were made to the nearest 1 cm. Weight (kg) was measured using a digital scale without shoes to the nearest 1 kg. Blood pressure (mm Hg) was measured in the right arm in sitting position using aneroid sphygmomanometer after 10 minutes of rest.

Venous blood samples were collected from all the participants after an overnight fast (of at least 8 hours) under aseptic precautions for quantifying the biochemical parameters (in mg/dL), including fasting blood glucose (FBG), total cholesterol (TC), serum triglycerides (TG), and high density lipoprotein (HDL-C). All biochemical assays were carried out using automated biochemical analyzers [Roche Cobas 6000 Chemistry Analyzer (Roche Diagnostics, Indianapolis, IN, U.S.A.)]. For conversion of units from mg/dL to mmol/L, TC and HDL-C values were divided by 38.67, and triglyceride values were divided by 88.57.

A positive history of diabetes mellitus and hypertension were considered based on self-reported history of illness or medications for the same, and/or laboratory/clinical measures suggestive of the condition – diabetes mellitus: FBG ≥ 126 mg/dL; hypertension: systolic/diastolic blood pressure ≥ 140/90 mm Hg. Cardiometabolic indices and risk scores were calculated from the data collected.

### 2.3. CALCULATION OF CARDIOMETABOLIC INDICES

A total of twenty cardiometabolic indices were computed from the demographic, anthropometric and biochemical data collected for every study participant. The indices computed and their formulae are given in *Table 1*. The units mentioned in the table footnote apply to the respective variables in every field of the table (unless mentioned otherwise).

**TABLE 1:**
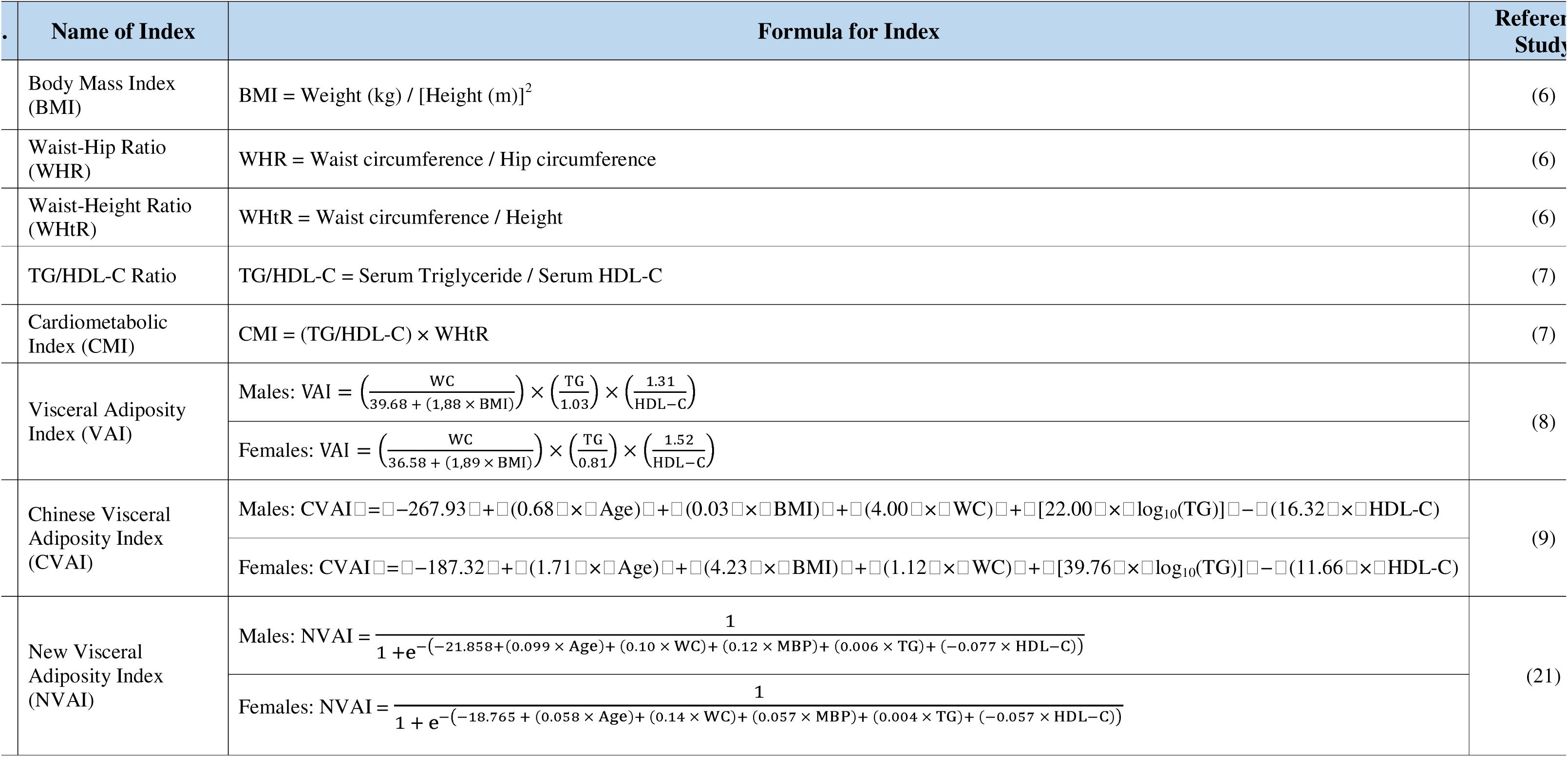
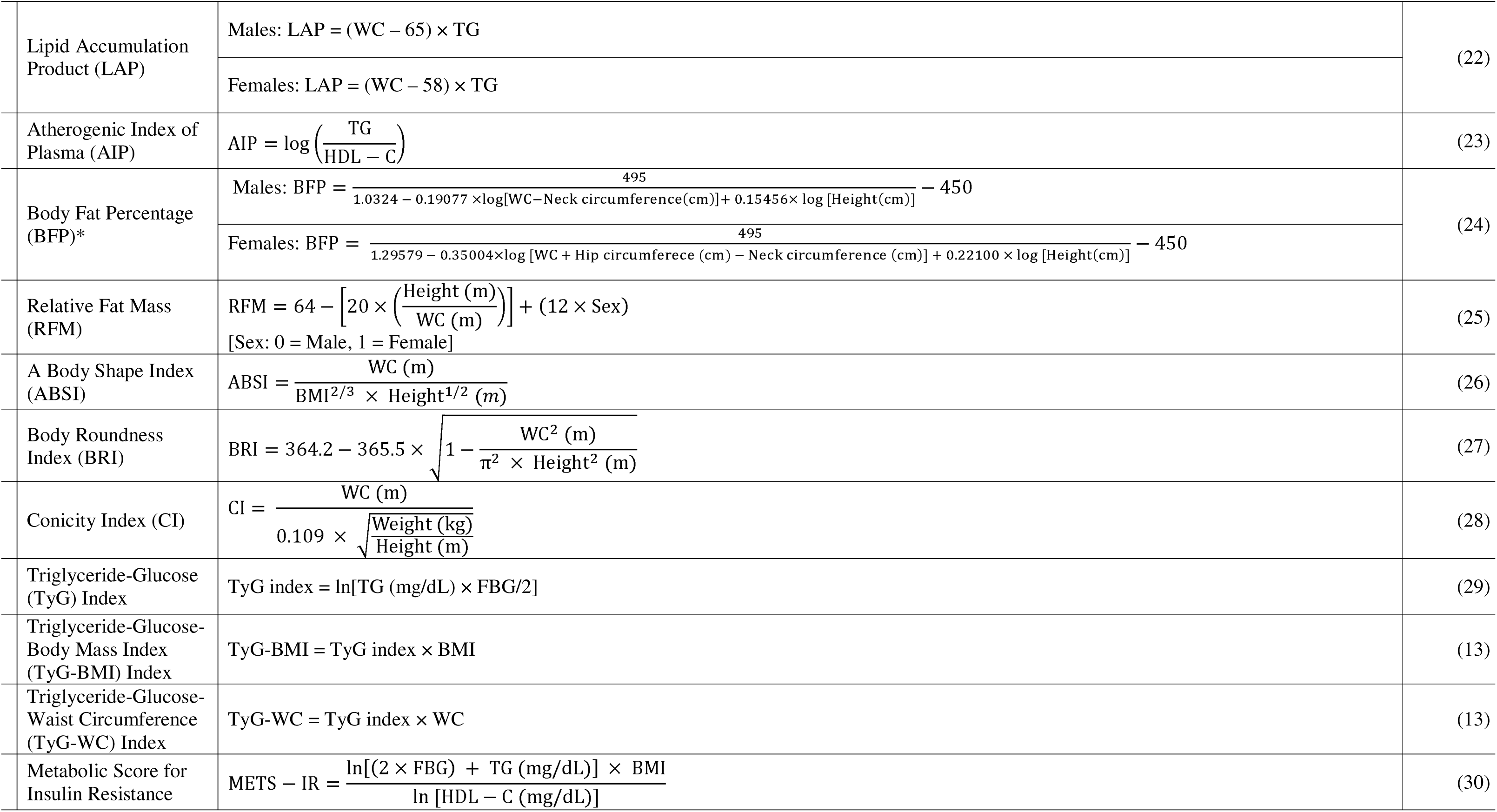
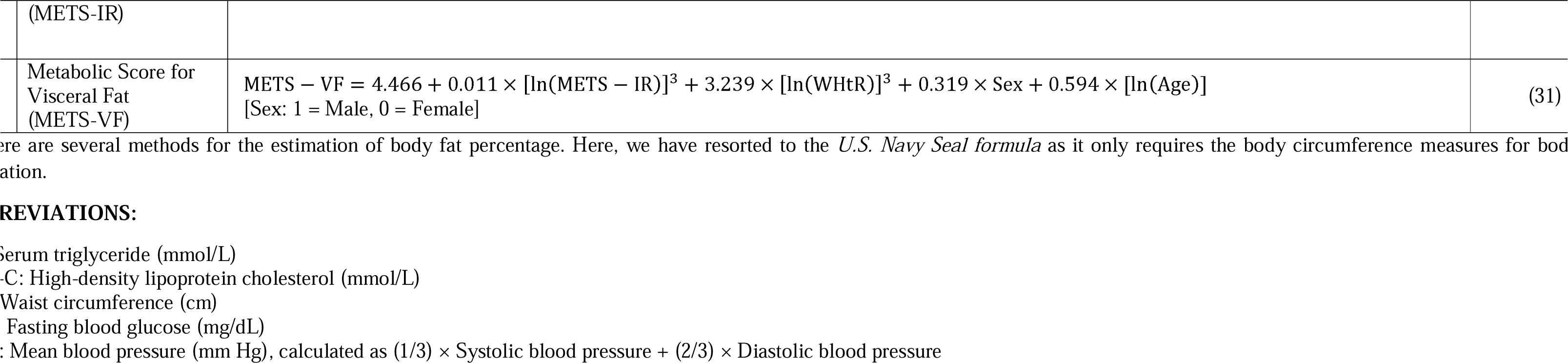
List of Cardiometabolic Indices analyzed in our study.

|  | Name of Index | Formula for Index | Referer Study |
| --- | --- | --- | --- |
| | Body Mass Index (BMI) | $BMI = \text{Weight (kg)} / [\text{Height (m)}]^2$ | (6) |
| | Waist-Hip Ratio (WHR) | $WHR = \text{Waist circumference} / \text{Hip circumference}$ | (6) |
| | Waist-Height Ratio (WHtR) | $WHtR = \text{Waist circumference} / \text{Height}$ | (6) |
| | TG/HDL-C Ratio | $TG/HDL-C = \text{Serum Triglyceride} / \text{Serum HDL-C}$ | (7) |
| | Cardiometabolic Index (CMI) | $CMI = (TG/HDL-C) \times WHtR$ | (7) |
| | Visceral Adiposity Index (VAI) | Males: $VAI = \left( \frac{WC}{39.68 + (1.88 \times BMI)} \right) \times \left( \frac{TG}{1.03} \right) \times \left( \frac{1.31}{HDL-C} \right)$ | (8) |
| | | Females: $VAI = \left( \frac{WC}{36.58 + (1.89 \times BMI)} \right) \times \left( \frac{TG}{0.81} \right) \times \left( \frac{1.52}{HDL-C} \right)$ | |
| | Chinese Visceral Adiposity Index (CVAI) | Males: $CVAI = -267.93 + (0.68 \times \text{Age}) + (0.03 \times \text{BMI}) + (4.00 \times \text{WC}) + [22.00 \times \log_{10}(TG)] - (16.32 \times \text{HDL-C})$ | (9) |
| | | Females: $CVAI = -187.32 + (1.71 \times \text{Age}) + (4.23 \times \text{BMI}) + (1.12 \times \text{WC}) + [39.76 \times \log_{10}(TG)] - (11.66 \times \text{HDL-C})$ | |
| | New Visceral Adiposity Index (NVAI) | Males: $NVAI = \frac{1}{1 + e^{-( -21.858 + (0.099 \times \text{Age}) + (0.10 \times \text{WC}) + (0.12 \times \text{MBP}) + (0.006 \times \text{TG}) + (-0.077 \times \text{HDL-C}))}}$ | (21) |
| | | Females: $NVAI = \frac{1}{1 + e^{-( -18.765 + (0.058 \times \text{Age}) + (0.14 \times \text{WC}) + (0.057 \times \text{MBP}) + (0.004 \times \text{TG}) + (-0.057 \times \text{HDL-C}))}}$ | |

|  |  |  |
| --- | --- | --- |
| Lipid Accumulation Product (LAP) | Males: $LAP = (WC - 65) \times TG$ | (22) |
| | Females: $LAP = (WC - 58) \times TG$ | |
| Atherogenic Index of Plasma (AIP) | $AIP = \log \left( \frac{TG}{HDL - C} \right)$ | (23) |
| Body Fat Percentage (BFP)* | Males: $BFP = \frac{495}{1.0324 - 0.19077 \times \log[WC - Neck\ circumference(cm)] + 0.15456 \times \log[Height(cm)]} - 450$ | (24) |
| | Females: $BFP = \frac{495}{1.29579 - 0.35004 \times \log[WC + Hip\ circumferece\ (cm) - Neck\ circumference\ (cm)] + 0.22100 \times \log[Height(cm)]} - 450$ | |
| Relative Fat Mass (RFM) | $RFM = 64 - \left[ 20 \times \left( \frac{Height\ (m)}{WC\ (m)} \right) \right] + (12 \times Sex)$<br>[Sex: 0 = Male, 1 = Female] | (25) |
| A Body Shape Index (ABSI) | $ABSI = \frac{WC\ (m)}{BMI^{2/3} \times Height^{1/2}\ (m)}$ | (26) |
| Body Roundness Index (BRI) | $BRI = 364.2 - 365.5 \times \sqrt{1 - \frac{WC^2\ (m)}{\pi^2 \times Height^2\ (m)}}$ | (27) |
| Conicity Index (CI) | $CI = \frac{WC\ (m)}{0.109 \times \sqrt{\frac{Weight\ (kg)}{Height\ (m)}}}$ | (28) |
| Triglyceride-Glucose (TyG) Index | $TyG\ index = \ln[TG\ (mg/dL) \times FBG/2]$ | (29) |
| Triglyceride-Glucose-Body Mass Index (TyG-BMI) Index | $TyG-BMI = TyG\ index \times BMI$ | (13) |
| Triglyceride-Glucose-Waist Circumference (TyG-WC) Index | $TyG-WC = TyG\ index \times WC$ | (13) |
| Metabolic Score for Insulin Resistance | $METS - IR = \frac{\ln[(2 \times FBG) + TG\ (mg/dL)] \times BMI}{\ln[HDL - C\ (mg/dL)]}$ | (30) |
| (METS-IR) |  |  |
| Metabolic Score for Visceral Fat (METS-VF) | $\text{METS} - \text{VF} = 4.466 + 0.011 \times [\ln(\text{METS} - \text{IR})]^3 + 3.239 \times [\ln(\text{WHtR})]^3 + 0.319 \times \text{Sex} + 0.594 \times [\ln(\text{Age})]$ [Sex: 1 = Male, 0 = Female] | (31) |

There are several methods for the estimation of body fat percentage. Here, we have resorted to the *U.S. Navy Seal formula* as it only requires the body circumference measures for body fat estimation.

### 2.4. CALCULATION OF RISK SCORES

#### (I) Framingham Risk Score (for Cardiovascular Risk)

The Framingham Risk Score (FRS) is a sex-specific risk stratification tool that is used to predict the 10-year cardiovascular risk in an individual. This tool is a product of the landmark Framingham Heart Study (FHS), the long-term cardiovascular cohort study in Massachusetts, U.S.A. In our study, we have employed the FHS 10-year cardiovascular disease risk assessment tool proposed by *D’Agostino RB Sr et al.* (32), specifically the lipid-based model using total cholesterol and HDL-C levels. This tool has been shown to possess a good predictive potential for cardiovascular risk in the Asian population (33).

This tool is applicable only to the individuals in the age group of 30 to 74 years without a history of cardiovascular disease at baseline assessment. Participants were stratified into low-risk (<10% of 10-year risk), intermediate-risk (10 – 20%), and high-risk (≥20%) groups based on their FRS. Since previous histories of cardiovascular disease, stroke, peripheral vascular diseases, and chronic kidney disease are coronary heart disease risk equivalents (according to NCEP ATP III), such patients have been categorized into the high-risk category (≥20% of 10-year risk) in our study.

#### (II) CHA_2_DS_2_-VASc Score (for Stroke Risk)

The CHA_2_DS_2_-VASc score is a point-based scoring scheme to ascertain the risk of stroke in patients with non-valvular atrial fibrillation (34). Although its primary utility is to guide the need for anticoagulation therapy in atrial fibrillation patients, the recent literature has shown that this tool can be used as a predictor of stroke risk even in patients without atrial fibrillation (35). Participants were categorized based on their total CHA_2_DS_2_-VASc score into low-risk (score = 0), medium-risk (score = 1), and high-risk (score ≥ 2) strata.

#### (III) Padua Prediction Score (for Risk of Venous Thromboembolism)

The Padua Prediction Score is a risk-prediction tool developed to predict the risk of venous thromboembolism in non-surgical hospitalized patients and determine the need for pharmacological prophylaxis (36). In our study, we have employed this tool for assessing the future risk of venous thromboembolism among the study participants. A score of ≥ 4 has considered to be the high-risk category, while those with a score < 4 were assigned to the low-risk category.

**Note:** Refer the Supplementary Materials section for the detailed risk score schemes

### 2.5. STATISTICAL ANALYSIS

Data collection was done on Microsoft Excel 2016 (Microsoft Corporation, Redmond, Washington, U.S.A.). Statistical analysis was performed on IBM SPSS Statistics for Windows, Version 26.0 (IBM Corporation, Armonk, NY, U.S.A.) and MedCalc Statistical Software, Version 22.023 (MedCalc Software Ltd, Ostend, Belgium).

Assessment of the normality of continuous data was done with Kolmogorov-Smirnov test. Continuous variables were presented as median (interquartile range), while categorical variables were expressed in frequencies and percentages.

Areas under the receiver operating characteristic (AUC) were used to assess the predictive efficacy of the cardiometabolic indices. Indices with AUC > 0.5 and p-value < 0.05 were regarded to have statistically significant predictive association with future thrombotic risk and considered for further analysis. All significant indicators were compared with other significant indicators to compare their predictive ability under each risk score. The indices with no significant difference (p-value > 0.05) were taken as possible best predictive indicators under each risk score. The indicator with the highest AUC was presented as the best predictor of the corresponding risk. Optimal cut-off points were identified as the point with the highest Youden’s index [J_max_ = maximum (sensitivity + specificity – 1)], as identified by MedCalc software (MedCalc Software Ltd, Ostend, Belgium).

Out of the indices displaying significant predictive ability for all risk scores, indices with no significant difference in predictive ability under each risk score were identified as probable candidates for the best overall thrombotic risk indicator. Optimal cut-off points for overall thrombotic risk was identified using the indicator value with the highest Youden’s index across all the three risk scores. However, since this value was not the same for all the risk scores in our case, the index value with a higher sensitivity for the considered risks was chosen. Statistical significance was accepted at 0.05 level (two-sided) and confidence interval (CI) was ensured at 95% level.

### 2.6. CONFIDENTIALITY

Confidentiality of the participants’ details was strictly maintained during this study. No names would be mentioned during publication. The participants were allotted a unique study reference number during data collection, and only this reference number was used during the analysis.

## 3. RESULTS

### 3.1. DEMOGRAPHIC CHARACTERISTICS OF THE STUDY POPULATION

A total of 200 participants were included in the study, out of which 117 (58.5%) were males and 83 (41.5%) were females. The median age was 50 (42–60) years. The baseline characteristics of the study population are given in *Table 2*. Out of the participants, 9.5%, 6% and 6% had a previous history of coronary artery disease, stroke, and venous thromboembolism, respectively. High-risk status in the Framingham, CHA_2_DS_2_-VASc and Padua prediction risk scores were significantly associated with the occurrence of these vascular complications (p-value < 0.05). Individuals in the high-risk scoring strata were more likely older, had a higher prevalence of diabetes and hypertension, and had raised levels of fasting blood glucose, HDL-C, and cardiometabolic indices, as compared to the non-high risk groups (p-value < 0.05).

**TABLE 2:** Baseline characteristics of the study population (n = 200)

| Parameters | Overall (n = 200)<br>[median (IQR)] | Males (n = 117)<br>[median (IQR)] | Females (n = 83)<br>[median (IQR)] |
| --- | --- | --- | --- |
| Age (years) | 50 (42 - 60) | 49 (40 - 59) | 50 (43 - 60) |
| Height (cm) | 164 (156 - 170) | 168 (165 - 173) | 155 (150 - 160) |
| Weight (kg) | 70 (62 - 82) | 75 (64 - 85) | 68 (60 - 78) |
| Neck circumference (cm) | 39 (36 - 41) | 40 (38 - 42) | 36 (34 - 40) |
| Waist circumference (cm) | 98 (88 - 106) | 98 (89 - 107) | 98 (87 - 105) |
| Hip circumference (cm) | 102 (93 - 110) | 100 (91 - 108) | 104 (97 - 114) |
| Systolic Blood Pressure (mm Hg) | 130 (120 - 140) | 130 (120 - 140) | 130 (120 - 140) |
| Diastolic Blood Pressure (mm Hg) | 80 (70 - 90) | 80 (70 - 90) | 80 (70 - 80) |
| Fasting Blood Glucose (mg/dL) | 105.5 (95.5 - 152.0) | 104.0 (95.0 - 133.0) | 116.0 (99.0 - 169.0) |
| Total Cholesterol (mg/dL) | 184.5 (154.5 - 209.0) | 189.0 (158.0 - 209.0) | 167.0 (152.0 - 211.0) |
| HDL-C (mg/dL) | 53.0 (44.0 - 61.0) | 53.0 (45.0 - 60.0) | 51.5 (44.0 - 63.0) |
| Serum Triglycerides (mg/dL) | 132.5 (110.5 - 162.5) | 136.0 (115.0 - 175.0) | 127.0 (106.0 - 150.0) |
| **Comorbidity History, n (%)** |  |  |  |
| Diabetes mellitus | 115 (57.5%) | 65 (55.6%) | 50 (60.2%) |
| Hypertension | 78 (39.0%) | 48 (41.0%) | 30 (36.1%) |
| Smoking | 37 (18.5%) | 37 (31.6%) | 0 (0.0%) |
| Previous coronary artery disease | 19 (9.5%) | 12 (10.3%) | 7 (8.4%) |
| Previous stroke | 12 (6.0%) | 10 (8.5%) | 2 (2.4%) |
| Previous venous thromboembolism | 12 (6.0%) | 4 (3.4%) | 8 (9.6%) |
| Chronic kidney disease | 26 (13.0%) | 16 (13.7%) | 10 (12%) |
| **Cardiometabolic Indices** |  |  |  |
| BMI | 26.79 (22.92 - 31.41) | 25.85 (22.68 - 29.75) | 28.67 (23.24 - 32.02) |
| WHR | 0.96 (0.93 - 1.00) | 0.98 (0.95 - 1.00) | 0.93 (0.87 - 0.97) |
| WHtR | 0.60 (0.54 - 0.66) | 0.58 (0.53 - 0.63) | 0.62 (0.55 - 0.69) |
| TG/HDL-C | 2.60 (2.04 - 3.61) | 2.65 (2.15 - 3.68) | 2.36 (1.86 - 3.44) |
| CMI | 1.51 (1.20 - 2.17) | 1.52 (1.23 - 2.13) | 1.47 (1.15 - 2.19) |
| VAI | 1.78 (1.38 - 2.62) | 1.62 (1.30 - 2.27) | 2.21 (1.54 - 2.99) |
| CVAI | 136.22 (94.29 - 164.43) | 144.44 (96.35 - 178.92) | 118.87 (88.82 - 149.59) |
| NVAI | 0.98 (0.89 - 1.00) | 0.99 (0.94 - 1.00) | 0.97 (0.83 - 0.99) |
| LAP | 56.36 (38.26 - 76.07) | 53.49 (37.7 - 76.21) | 58.17 (38.61 - 75.60) |
| AIP | 0.41 (0.31 - 0.56) | 0.42 (0.33 - 0.57) | 0.37 (0.27 - 0.54) |
| BFP | 30.62 (24.29 - 41.17) | 26.38 (20.43 - 30.35) | 43.07 (36.71 - 48.86) |
| RFM | 33.26 (28.70 - 42.74) | 29.80 (25.94 - 32.35) | 43.96 (39.78 - 47.11) |
| ABSI | 0.08 (0.08 - 0.09) | 0.08 (0.08 - 0.09) | 0.08 (0.08 - 0.09) |
| BRI | 5.37 (4.07 - 6.82) | 5.09 (3.85 - 6.17) | 5.99 (4.39 - 7.69) |
| CI | 1.35 (1.29 - 1.40) | 1.35 (1.29 - 1.40) | 1.34 (1.29 - 1.42) |
| TyG Index | 8.93 (8.68 - 9.33) | 8.93 (8.74 - 9.29) | 8.95 (8.61 - 9.46) |
| TyG-BMI | 243.55 (205.81 - 282.71) | 234.98 (206.47 - 269.00) | 258.96 (205.32 - 293.32) |
| TyG-WC | 896.13 (788.55 - 972.66) | 898.85 (798.91 - 969.67) | 881.42 (775.06 - 975.92) |
| METS-IR | 40.35 (34.89 - 47.65) | 39.04 (33.83 - 44.91) | 42.94 (36.70 - 49.08) |
| METS-VF | 7.53 (7.36 - 7.70) | 7.66 (7.51 - 7.78) | 7.38 (7.25 - 7.50) |
| **Risk Scores** |  |  |  |
| **Framingham Risk Score** | 13 (8 - 17) | 13 (8 - 17) | 12 (7 - 17) |
| High risk, n (%) | 87 (43.5%) | 60 (51.3%) | 27 (32.5%) |
| Intermediate risk, n (%) | 36 (18.0%) | 17 (14.5%) | 19 (22.9%) |
| Low risk, n (%) | 77 (38.5%) | 40 (34.2%) | 37 (44.6%) |
| **CHA<sub>2</sub>DS<sub>2</sub>-VASc Score** | 2 (1 - 3) | 1 (0 - 2) | 2 (1 - 3) |
| High risk, n (%) | 101 (50.5%) | 40 (34.2%) | 61 (73.5%) |
| Intermediate risk, n (%) | 65 (32.5%) | 43 (36.8%) | 22 (26.5%) |
| Low risk, n (%) | 34 (17.0%) | 34 (29.1%) | 0 (0.0%) |
| **Padua Prediction Score** | 1 (0 - 4) | 1 (0 - 3) | 2 (1 - 4) |
| High risk, n (%) | 50 (25.0%) | 24 (20.5%) | 26 (31.3%) |
| Low risk, n (%) | 150 (75.0%) | 93 (79.5%) | 57 (68.7%) |

**Note:** Results have been expressed as Median (Interquartile range) for continuous variables, or as Number (Percentage) for categorical variables.
**Abbreviations:** HDL-C: high-density lipoprotein cholesterol, BMI: Body Mass Index, WHR: Waist-Hip Ratio, WHtR: Waist-Height Ratio, TG/HDL-C: TG/HDL-C Ratio, CMI: Cardiometabolic Index, VAI: Visceral Adiposity Index, CVAI: Chinese Visceral Adiposity Index, NVAI: New Visceral Adiposity Index, LAP: Lipid Accumulation Product, AIP: Atherogenic Index of Plasma, BFP: Body Fat Percentage, RFM: Relative Fat Mass, ABSI: A Body Shape Index, BRI: Body Roundness Index, CI: Conicity Index, TyG Index: Triglyceride-Glucose Index, TyG-BMI: Triglyceride-Glucose-Body Mass Index Index, TyG-WC: Triglyceride-Glucose-Waist Circumference Index, METS-IR: Metabolic Score for Insulin Resistance, METS-VF: Metabolic Score for Visceral Fat.

### 3.2. RECEIVER OPERATING CHARACTERISTIC (ROC) CURVE ANALYSIS

Receiver-operating characteristic curve analysis was undertaken to identify cardiometabolic indices which possess a significant predictive value for the thrombotic complications (*Table 3, Figures 1, 2* and *3*). All indices with area under ROC curve (AUC) greater than 0.5 and p-value < 0.05 were considered to have statistically significant predictive potential and were subjected to further analysis. Eight indices for FRS, eleven indices for CHA_2_DS_2_-VASc score, and none of the indices for Padua prediction score were found to meet the criteria.

**FIGURE 1:**
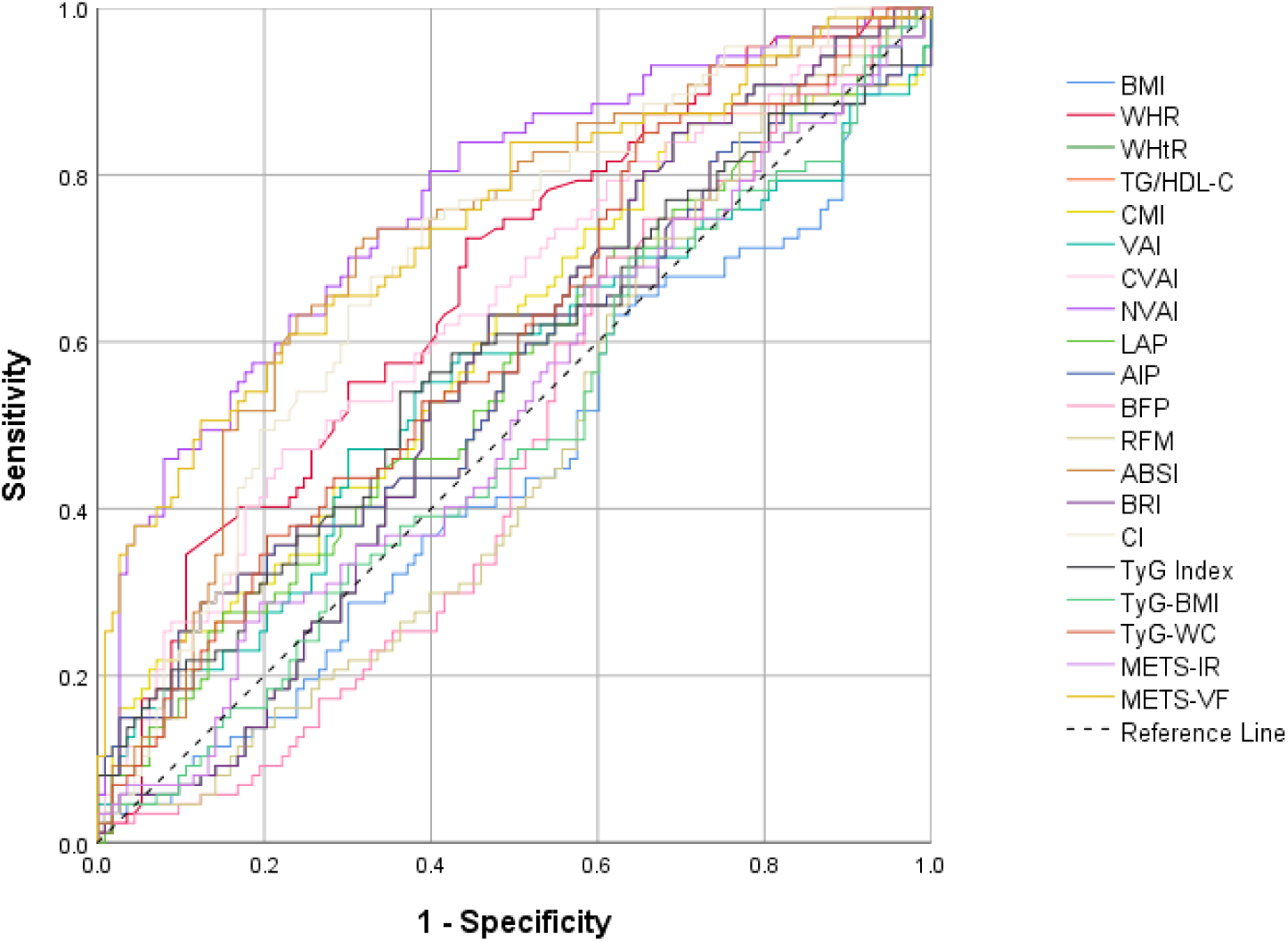
ROC Curve for Framingham Risk Score **Note:** Significant predictive value (AUC > 0.5 and p-value < 0.05) has been observed with NVAI, METS-VF, ABSI, CI, WHR, CVAI, TyG-WC, and CMI (in decreasing order of AUCs)

**FIGURE 2:**
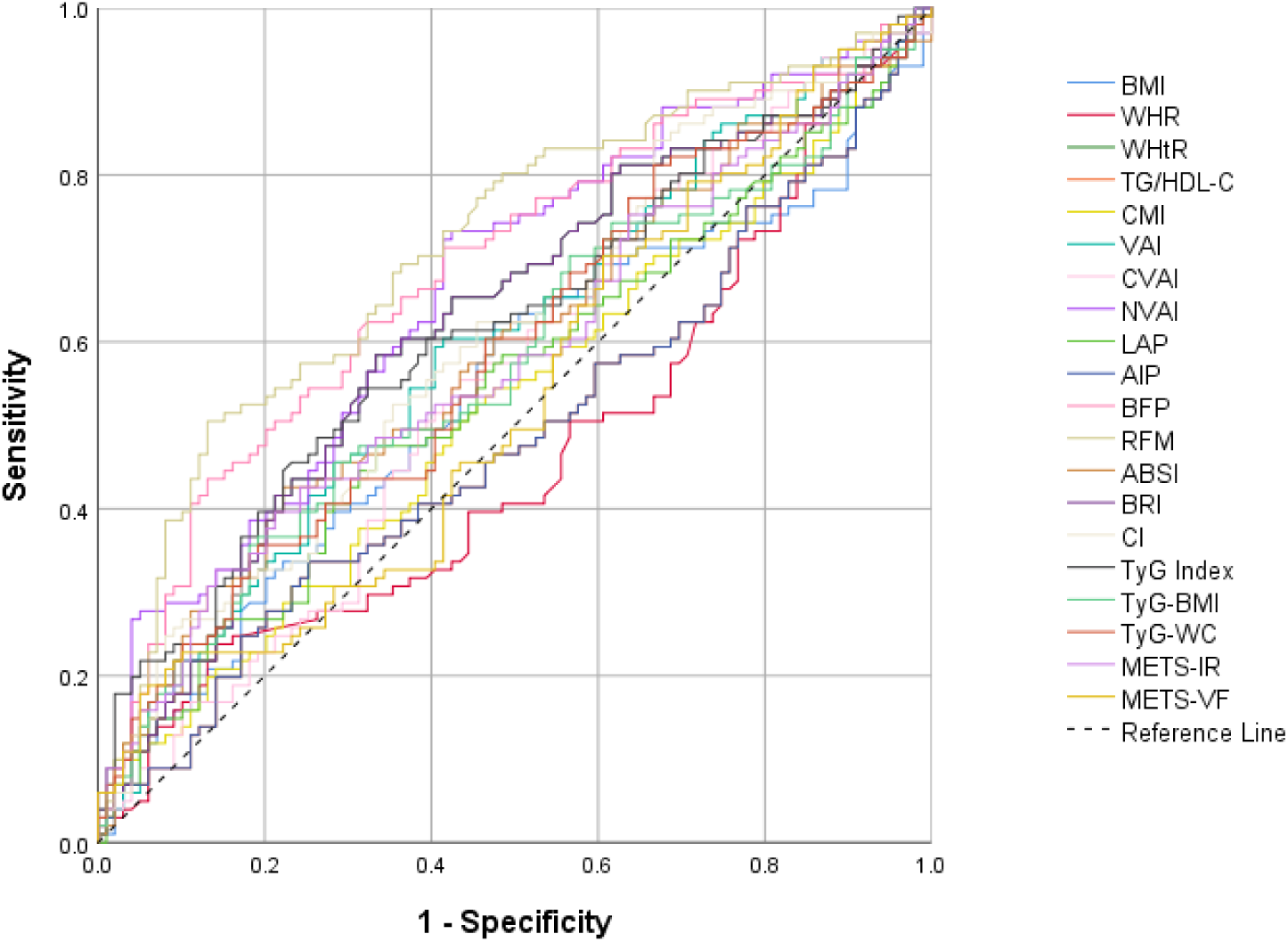
ROC Curve for CHA_2_DS_2_VASc Score **Note:** Significant predictive value (AUC > 0.5 and p-value < 0.05) has been observed with RFM, BFP, NVAI, WHtR, BRI, TyG-index, CI, VAI, ABSI, TyG-WC, and METS-IR (in decreasing order of AUCs)

**FIGURE 3:**
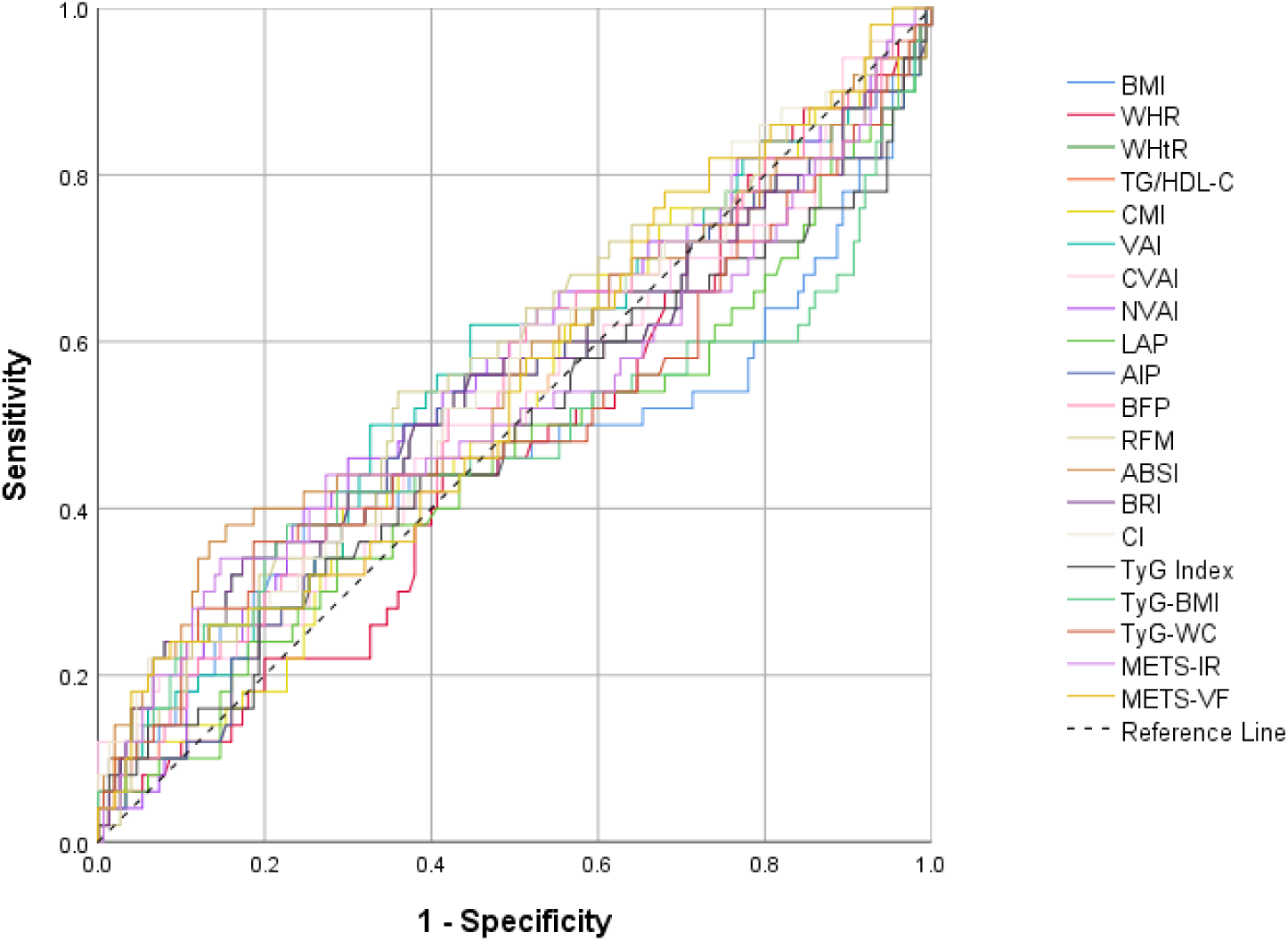
ROC Curve for Padua Prediction Score **Note:** None of the indices have displayed significant predictive value (AUC > 0.5 and p-value < 0.05)

**TABLE 3:** Receiver Operating Characteristic analysis of indices across risk scores.

| S.No. | Cardiometabolic Indices | Framingham Risk Score |  | CHA<sub>2</sub>DS<sub>2</sub>-VASc Score |  | Padua Prediction Score |  |
| --- | --- | --- | --- | --- | --- | --- | --- |
|  |  | AUC (95% CI) | p-value | AUC (95% CI) | p-value | AUC (95% CI) | p-value |
| 1 | BMI | 0.547<br>(0.476 – 0.618) | 0.2533 | 0.542<br>(0.470 - 0.612) | 0.3124 | 0.535<br>(0.463 - 0.606) | 0.5255 |
| 2 | WHR | 0.668<br>(0.598 - 0.733) | **<0.0001** | 0.536<br>(0.464 - 0.606) | 0.3857 | 0.526<br>(0.455 - 0.597) | 0.5793 |
| 3 | WHtR | 0.556<br>(0.484 - 0.626) | 0.1677 | 0.622<br>(0.551 - 0.689) | **0.0022** | 0.542<br>(0.470 - 0.612) | 0.4239 |
| 4 | TG/HDL-C | 0.560<br>(0.488 - 0.630) | 0.1557 | 0.514<br>(0.443 - 0.585) | 0.7314 | 0.522<br>(0.450 - 0.593) | 0.6660 |
| 5 | CMI | 0.596<br>(0.524 - 0.664) | **0.0196** | 0.523<br>(0.452 - 0.594) | 0.5711 | 0.521<br>(0.450 - 0.592) | 0.6597 |
| 6 | VAI | 0.550<br>(0.478 - 0.620) | 0.2402 | 0.595<br>(0.523 - 0.663) | **0.0186** | 0.555<br>(0.483 - 0.625) | 0.2729 |
| 7 | CVAI | 0.641<br>(0.570 - 0.707) | **0.0004** | 0.554<br>(0.483 - 0.624) | 0.1841 | 0.522<br>(0.450 - 0.593) | 0.6688 |
| 8 | NVAI | 0.769<br>(0.704 - 0.825) | **<0.0001** | 0.663<br>(0.593 - 0.728) | **<0.0001** | 0.558<br>(0.486 - 0.628) | 0.2398 |
| 9 | LAP | 0.556<br>(0.484 - 0.626) | 0.1750 | 0.545<br>(0.473 - 0.615) | 0.2768 | 0.543<br>(0.471 - 0.614) | 0.4000 |
| 10 | AIP | 0.560<br>(0.488 - 0.630) | 0.1557 | 0.514<br>(0.443 - 0.585) | 0.7314 | 0.522<br>(0.450 - 0.593) | 0.6660 |
| 11 | BFP | 0.531<br>(0.459 - 0.601) | 0.4547 | 0.684<br>(0.615 - 0.748) | **<0.0001** | 0.543<br>(0.472 - 0.614) | 0.3807 |
| 12 | RFM | 0.532<br>(0.460 - 0.603) | 0.4354 | 0.714<br>(0.646 - 0.776) | **<0.0001** | 0.566<br>(0.494 - 0.636) | 0.1797 |
| 13 | ABSI | 0.717<br>(0.650 - 0.779) | **<0.0001** | 0.589<br>(0.518 - 0.658) | **0.0273** | 0.565<br>(0.493 - 0.634) | 0.2158 |
| 14 | BRI | 0.556<br>(0.484 - 0.626) | 0.1677 | 0.622<br>(0.551 - 0.689) | **0.0022** | 0.542<br>(0.470 - 0.612) | 0.4239 |
| 15 | CI | 0.698<br>(0.629 - 0.761) | **<0.0001** | 0.604<br>(0.532 - 0.672) | **0.0095** | 0.559<br>(0.487 - 0.629) | 0.2368 |
| 16 | TyG Index | 0.576<br>(0.504 - 0.645) | 0.0678 | 0.617<br>(0.546 - 0.685) | **0.0032** | 0.516<br>(0.444 - 0.587) | 0.7626 |
| 17 | TyG-BMI | 0.513<br>(0.442 - 0.584) | 0.7508 | 0.573<br>(0.501 - 0.642) | 0.0745 | 0.525<br>(0.453 - 0.596) | 0.6590 |
| 18 | TyG-WC | 0.599<br>(0.528 - 0.668) | **0.0138** | 0.584<br>(0.512 - 0.653) | **0.0375** | 0.504<br>(0.432 - 0.575) | 0.9472 |
| 19 | METS-IR | 0.514<br>(0.443 - 0.585) | 0.7307 | 0.581<br>(0.510 - 0.651) | **0.0447** | 0.525<br>(0.454 - 0.596) | 0.6359 |
| 20 | METS-VF | 0.744<br>(0.678 - 0.803) | **<0.0001** | 0.533<br>(0.461 - 0.604) | 0.4244 | 0.548<br>(0.476 - 0.618) | 0.3281 |

**Note:** Statistically significant p-values (<0.05) have been given in bold for the ease of the readers Null hypothesis for the ROC analysis is AUC = 0.5
**Abbreviations:** AUC: Area under the ROC curve; 95% CI: 95% confidence interval, BMI: Body Mass Index, WHR: Waist-Hip Ratio, WHtR: Waist-Height Ratio, TG/HDL-C: TG/HDL-C Ratio, CMI: Cardiometabolic Index, VAI: Visceral Adiposity Index, CVAI: Chinese Visceral Adiposity Index, NVAI: New Visceral Adiposity Index, LAP: Lipid Accumulation Product, AIP: Atherogenic Index of Plasma, BFP: Body Fat Percentage, RFM: Relative Fat Mass, ABSI: A Body Shape Index, BRI: Body Roundness Index, CI: Conicity Index, TyG Index: Triglyceride-Glucose Index, TyG-BMI: Triglyceride-Glucose-Body Mass Index Index, TyG-WC: Triglyceride-Glucose-Waist Circumference Index, METS-IR: Metabolic Score for Insulin Resistance, METS-VF: Metabolic Score for Visceral Fat.

#### 3.2.1. Framingham Risk Score

Among the twenty indices, NVAI, METS-VF, ABSI, CI, WHR, CVAI, TyG-WC, and CMI (in decreasing order of AUCs) have shown significant predictive potential for coronary artery disease. *Table 4* gives the optimal cut-off points with the corresponding sensitivity, specificity and Youden index for the significant indices. NVAI has the highest AUC value in predicting 10-year cardiovascular risk (AUC = 0.769, 95% CI = 0.704 - 0.825), followed by METS-VF (AUC = 0.744, 95% CI = 0.678 - 0.803) and ABSI (AUC = 0.717, 95% CI = 0.650 - 0.779). The predictive performance of NVAI was significantly better than other indices (p-value < 0.05) as assessed by DeLong’s method, except for ABSI (p-value = 0.301), CI (p-value = 0.158) and METS-VF (p value = 0.614). A significant gender variation in AUC was not observed among the eight indices, except for NVAI (p-value = 0.022; male cut-off = 0.9932, female cut-off = 0.9634).

**TABLE 4:** Indices with significant predictive value for Framingham risk score.

| S.No. | Cardiometabolic Indices | Optimal Cut-off Points | Sensitivity (%) | Specificity (%) | Youden Index, $J_{\max}$ |
| --- | --- | --- | --- | --- | --- |
| 1 | NVAI | 0.972 | 80.46 | 60.18 | 0.4064 |
| 2 | METS-VF | 7.677 | 50.57 | 87.61 | 0.3819 |
| 3 | ABSI | 0.085 | 72.41 | 68.14 | 0.4056 |
| 4 | CI | 1.340 | 74.71 | 61.06 | 0.3577 |
| 5 | WHR | 0.955 | 72.41 | 55.75 | 0.2817 |
| 6 | CVAI | 152.073 | 47.13 | 77.88 | 0.2500 |
| 7 | TyG-WC | 790.257 | 85.06 | 34.51 | 0.1957 |
| 8 | CMI | 1.196 | 83.91 | 31.86 | 0.1577 |

**Abbreviations:** NVAI: New Visceral Adiposity Index, METS-VF: Metabolic Score for Visceral Fat, ABSI: A Body Shape Index, CI: Conicity Index, WHR: Waist-Hip Ratio, CVAI: Chinese Visceral Adiposity Index, TyG-WC: Triglyceride-Glucose-Waist Circumference Index, CMI: Cardiometabolic Index

#### 3.2.2. CHA_2_DS_2_VASc Score

From the indices evaluated, RFM, BFP, NVAI, WHtR, BRI, TyG-index, CI, VAI, ABSI, TyG-WC, and METS-IR (in decreasing order of AUCs) have demonstrated statistically significant predictive association with CHA_2_DS_2_VASc Score. *Table 5* shows the indices with predictive value for stroke risk and their optimal cut-off points, sensitivity, specificity and Youden index. On ROC analysis, RFM has the highest predictive value for stroke (AUC = 0.714, 95% CI = 0.646 - 0.776), followed by BFP (AUC = 0.684, 95% CI = 0.615 - 0.748) and NVAI (AUC = 0.663, 95% CI = 0.593 - 0.728). RFM has significantly higher predictive value as compared to the other indices (p-value < 0.05, DeLong’s method), except for BFP (p-value = 0.572), NVAI (p-value = 0.336), WHtR (p-value = 0.091), BRI (p-value = 0.091), and TyG index (p-value = 0.075). A significant gender-wise split was noticed only with TyG index (p-value = 0.003) with a male cut-off = 9.8274 and female cut-off = 8.704.

**TABLE 5:** Indices with significant predictive value for CHA_2_DS_2_VASc score.

| S.No. | Cardiometabolic Indices | Optimal Cut-off Points | Sensitivity (%) | Specificity (%) | Youden Index, J<sub>max</sub> |
| --- | --- | --- | --- | --- | --- |
| 1 | RFM | 39.182 | 50.50 | 86.87 | 0.3736 |
| 2 | BFP | 38.466 | 43.56 | 86.87 | 0.3043 |
| 3 | NVAI | 0.974 | 72.28 | 58.59 | 0.3086 |
| 4 | WHtR | 0.607 | 58.42 | 66.67 | 0.2508 |
| 5 | BRI | 5.586 | 58.42 | 66.67 | 0.2508 |
| 6 | TyG Index | 9.147 | 44.55 | 77.78 | 0.2233 |
| 7 | CI | 1.286 | 84.16 | 33.33 | 0.1749 |
| 8 | VAI | 1.773 | 59.41 | 59.60 | 0.1900 |
| 9 | ABSI | 0.087 | 42.57 | 77.78 | 0.2035 |
| 10 | TyG-WC | 969.667 | 34.65 | 81.82 | 0.1647 |
| 11 | METS-IR | 47.873 | 32.67 | 85.86 | 0.1853 |

**Abbreviations:** RFM: Relative Fat Mass, BFP: Body Fat Percentage, NVAI: New Visceral Adiposity Index, WHtR: Waist-Height Ratio, BRI: Body Roundness Index, TyG Index: Triglyceride-Glucose Index, CI: Conicity Index, VAI: Visceral Adiposity Index, ABSI: A Body Shape Index, TyG-WC: Triglyceride-Glucose-Waist Circumference Index, METS-IR: Metabolic Score for Insulin Resistance

#### 3.2.3. Padua Prediction Score

On examining our list of cardiometabolic indices, none of the indices were found to possess a p-value < 0.05 and AUC > 0.5 for Padua prediction score, and thus were not found to bear any statistically significant predictive potential for venous thromboembolism.

#### 3.2.4. Best Overall Predictors of Thrombotic Catastrophe

Since no significant indices were found with Padua prediction score, our best overall predictor shall be restricted only to the FRS and CHA_2_DS_2_VASc scores. Among the twenty indices, only four cardiometabolic indices have displayed statistical significance for both risk scores, namely – NVAI, ABSI, CI, and TyG-WC.

Out of these indices, NVAI possesses the highest AUC for FRS (AUC = 0.769, 95% CI = 0.704 - 0.825), as well as a non-significant difference (p-value = 0.336) in stroke predictive potential when compared against RFM (which has the highest AUC under CHA_2_DS_2_VASc score). In contrast to this, neither ABSI, CI, nor TyG-WC has a statistically non-significant difference in predictive potential when compared against the best individual risk predictors (indices with the highest AUC under each risk score, namely – NVAI for FRS, and RFM for CHA_2_DS_2_VASc score). This makes NVAI the best overall predictor of both cardiovascular and stroke risks.

The optimal cut-off for NVAI for overall prediction of coronary and stroke risks was found out to be at 0.972 (FRS: sensitivity = 80.5%, specificity = 60.2%; CHA_2_DS_2_VASc score: sensitivity = 72.3%, specificity = 57.6%). A gender-specific cut-off has also been attempted in our study – 0.993 for males (FRS: sensitivity = 70.0%, specificity = 80.7%; CHA_2_DS_2_VASc score: sensitivity = 72.5%, specificity = 70.2%), and 0.913 for females (FRS: sensitivity = 74.1%, specificity = 41.1%; CHA_2_DS_2_VASc score: sensitivity = 73.8%, specificity = 68.2%). Values of NVAI greater than or equal to these cut-off points correspond to the high risk strata for both cardiovascular and cerebrovascular events in the near future.

## 4. DISCUSSION

Thrombovascular events such as ischaemic heart disease, stroke and venous thromboembolism are leading causes of mortality worldwide and there has been an ever-rising effort towards their screening and early detection. Cardiometabolic indices have been utilized as reliable and convenient tools for identifying high-risk individuals in the population based on their risk factor profile. Conventionally, body mass index (BMI) and waist circumference (WC) have been preferred by practitioners, although several novel indices have been discussed in the literature over the years. Previous studies which have assessed the discriminatory potential of such indices have shown that the optimal choice of the surrogate markers varies from one population to another (15,16). Indians and South Asians possess a ‘thin-fat’ phenotype with greater visceral adiposity and more insulin resistance as compared to their western counterparts (37,38). Our study provides a population-specific comparative evidence on the predictive potential of conventional and novel cardiometabolic indices for the Indian demography.

Our study is a single-centre, hospital-based cross-sectional study on 200 participants (median age = 50 years, males:females = 1.4:1) visiting the master health check-up centre of our tertiary care institution in India. Out of the twenty indices evaluated, we have identified eight indices (namely, NVAI, METS-VF, ABSI, CI, WHR, CVAI, TyG-WC, and CMI) that have shown significant discriminatory potential for 10-year cardiovascular disease risk, and eleven indices (namely, RFM, BFP, NVAI, WHtR, BRI, TyG-index, CI, VAI, ABSI, TyG-WC, and METS-IR) that have displayed predictive potential for stroke risk (in decreasing order of AUCs). Four indices (namely, NVAI, ABSI, CI, and TyG-WC) are statistically significant predictors of both cardiovascular and cerebrovascular events. Out of these indices, NVAI has turned out to be the best predictor of future thrombovascular catastrophe for the Indian demography (FRS AUC = 0.769, CHA_2_DS_2_VASc AUC = 0.663; p-value < 0.05).

The New Visceral Adiposity Index (NVAI) was developed as an indirect marker of cardiometabolic risk for the Korean population (Oh S-K et al., 2018), as a modification of the previous Visceral Adiposity Index (VAI) (21). It is calculated based on the age, waist circumference, mean blood pressure, TG, and HDL-C of the participants. It incorporates quantitative anthropometric measures of visceral adiposity as well as qualitative biochemical factors of adipose dysfunction. NVAI has shown significant correlation with atherosclerotic cardiovascular disease risk, coronary artery calcification, and renal dysfunction in the previous studies (10,21,39). Through our analysis on NVAI, we have arrived at an optimal cut-off point of 0.972 (males = 0.993, females = 0.913). Values of NVAI above this correspond to high-risk strata for coronary and stroke risks for the Indian population.

Apart from NVAI, other novel indices such as ABSI, CI and TyG-WC have displayed good predictive performance in our analysis. A Body Shape Index (ABSI), proposed by Krakauer et al. (2012) as an indicator of mortality hazard based on the NHANES database from the United States population, has been shown to be useful as surrogate marker for central obesity and cardiometabolic risk (11,26). Studies on the Conicity index (CI) have found significant correlation with cardiovascular risk and dyslipidemia, and has been shown to be an independent risk factor for all-cause mortality in non-cancerous elderly Chinese adults (12). Triglyceride-Glucose-Waist circumference (TyG-WC) index is a modification of TyG-Index which has shown predictive value for cardiovascular risk in the Korean population (40). Outcomes from our study are in line with the findings from previous studies on cardiovascular risk, and have additionally highlighted the ability of these indices to predict future stroke risk in the Indian population (AUC > 0.5 and p-value < 0.05 for CHA_2_DS_2_VASc score).

It is clear from our study that the commonly-used BMI is ineffective as a screening tool for thrombovascular events (p-value > 0.05 for both FRS and CHA_2_DS_2_VASc scores). This can be attributed to the fact that BMI fails to discriminate fat mass from lean mass, and subcutaneous adiposity from visceral adiposity, and does not factor into account the biochemical derangements associated with metabolic syndrome. This inadequacy emphasizes the need to transition towards more reliable indicators in screening programmes.

An interesting observation from our study is that the performance of cardiometabolic indices in predicting the risk for venous thromboembolism has been quite unsatisfactory. None of the indices evaluated in the study have displayed statistically significant predictive value for future thromboembolic events. This underscores the inherent difference in the risk factor profile for arterial and venous thrombosis, and highlights the limited utility of cardiometabolic indices in their prediction.

Through our study, we intend to modify the choice of cardiometabolic indices used for population screening. This demography-specific validation offers better thrombovascular risk prediction and can prove to quite helpful in identifying high-risk individuals from the masses. We have observed that indices ascertained by a combination of anthropometric measurements (quantitative measure) and biochemical parameters (qualitative measure) of visceral adipose dysfunction have displayed better predictive potential for future cardiovascular and cerebrovascular catastrophe over indices solely based on one of the measures. These indices could be used in large-scale non-communicable disease screening programmes, or as a quick tool for clinical risk assessment prior to making treatment decisions at the practitioner’s desk (for instance, for assessing the patient’s thrombotic risk before administering oral contraceptive pills).

Our study is however not without its own limitations. Small sample size, cross-sectional design, and the use of risk scoring schemes instead of actual incidence data are some shortcoming of our work. Future studies will be necessary for gaining external validation of our results and evaluating the outcomes on a long-term prospective manner. We also advocate future research efforts for exploring the possibility of increasing the screening sensitivity by using a combination of cardiometabolic indices for assessing the thrombovascular risk of the population.

## Data Availability

The data that support the findings of this study are available from the corresponding author upon reasonable request.

## 5. DECLARATIONS

The authors declare no conflicts of interest. The authors have not received funding from any sources for conducting this research work. All authors certify that they have no affiliations with or involvement in any organization or entity with any financial interest or non-financial interest in the subject matter or materials discussed in this manuscript.

Institutional ethics committee approval was obtained prior to the commencement of the study. Written informed consent was obtained from all participants prior to the interviews. The data that support the findings of this study are available from the corresponding author upon reasonable request. Clinical trial number: not applicable.

## 6. ACKNOWLEDGEMENTS

We wish to express our sincere gratitude to all participants who have taken part in this study. We would like to thank Dr. Gokulakrishnan, Assistant Professor, Institute of Internal Medicine, Madras Medical College, for helping us in collecting patients’ data at the Master Health Check-up centre of the hospital. We are grateful to the Institutes of Biochemistry and Pathology, Madras Medical College, for providing us with the laboratory facilities for carrying out the investigations needed for our study.

## REFERENCES

1. Canto JG, Kiefe CI, Rogers WJ, Peterson ED, Frederick PD, French WJ, et al. Number of coronary heart disease risk factors and mortality in patients with first myocardial infarction. JAMA. 2011 Nov 16;306(19):2120–7.

2. Rochlani Y, Pothineni NV, Kovelamudi S, Mehta JL. Metabolic syndrome: pathophysiology, management, and modulation by natural compounds. Therapeutic Advances in Cardiovascular Disease. 2017 Aug 1;11(8):215–25.

3. Vilahur G, Ben-Aicha S, Badimon L. New insights into the role of adipose tissue in thrombosis. Cardiovascular Research. 2017 Jul 1;113(9):1046–54.

4. Kwon H, Kim D, Kim JS. Body Fat Distribution and the Risk of Incident Metabolic Syndrome: A Longitudinal Cohort Study. Sci Rep. 2017 Sep 8;7(1):10955.

5. Shuster A, Patlas M, Pinthus JH, Mourtzakis M. The clinical importance of visceral adiposity: a critical review of methods for visceral adipose tissue analysis. British Journal of Radiology. 2012 Jan 1;85(1009):1–10.

6. Oumer A, Ale A, Tariku Z, Hamza A, Abera L, Seifu A. Waist-to-hip circumference and waist-to-height ratio could strongly predict glycemic control than body mass index among adult patients with diabetes in Ethiopia: ROC analysis. PLOS ONE. 2022 Nov 9;17(11):e0273786.

7. Shi WR, Wang HY, Chen S, Guo XF, Li Z, Sun YX. Estimate of prevalent diabetes from cardiometabolic index in general Chinese population: a community-based study. Lipids Health Dis. 2018 Oct 12;17(1):236.

8. Amato MC, Giordano C, Galia M, Criscimanna A, Vitabile S, Midiri M, et al. Visceral Adiposity Index: a reliable indicator of visceral fat function associated with cardiometabolic risk. Diabetes Care. 2010 Apr;33(4):920–2.

9. Xia MF, Chen Y, Lin HD, Ma H, Li XM, Aleteng Q, et al. A indicator of visceral adipose dysfunction to evaluate metabolic health in adult Chinese. Sci Rep. 2016 Dec 1;6:38214.

10. Son DH, Ha HS, Lee HS, Han D, Choi SY, Chun EJ, et al. Association of the new visceral adiposity index with coronary artery calcification and arterial stiffness in Korean population. Nutrition, Metabolism and Cardiovascular Diseases. 2021 Jun 7;31(6):1774– 81.

11. Bertoli S, Leone A, Krakauer NY, Bedogni G, Vanzulli A, Redaelli VI, et al. Association of Body Shape Index (ABSI) with cardio-metabolic risk factors: A cross-sectional study of 6081 Caucasian adults. PLOS ONE. 2017 Sep 25;12(9):e0185013.

12. Zhang A, Li Y, Ma S, Bao Q, Sun J, Cai S, et al. Conicity-index predicts all-cause mortality in Chinese older people: a 10-year community follow-up. BMC Geriatrics. 2022 Dec 16;22(1):971.

13. Lim J, Kim J, Koo SH, Kwon GC. Comparison of triglyceride glucose index, and related parameters to predict insulin resistance in Korean adults: An analysis of the 2007-2010 Korean National Health and Nutrition Examination Survey. PLoS One. 2019;14(3):e0212963.

14. Mazidi M, Kengne AP, Katsiki N, Mikhailidis DP, Banach M. Lipid accumulation product and triglycerides/glucose index are useful predictors of insulin resistance. Journal of Diabetes and its Complications. 2018 Mar 1;32(3):266–70.

15. Liu J, Tse LA, Liu Z, Rangarajan S, Hu B, Yin L, et al. Predictive Values of Anthropometric Measurements for Cardiometabolic Risk Factors and Cardiovascular Diseases Among 44 048 Chinese. J Am Heart Assoc. 2019 Aug 9;8(16):e010870.

16. Huang YC, Huang JC, Lin CI, Chien HH, Lin YY, Wang CL, et al. Comparison of Innovative and Traditional Cardiometabolic Indices in Estimating Atherosclerotic Cardiovascular Disease Risk in Adults. Diagnostics (Basel). 2021 Mar 28;11(4):603.

17. Krishnamoorthy Y, Rajaa S, Murali S, Rehman T, Sahoo J, Kar SS. Prevalence of metabolic syndrome among adult population in India: A systematic review and meta-analysis. PLOS ONE. 2020 Oct 19;15(10):e0240971.

18. Noubiap JJ, Nansseu JR, Lontchi-Yimagou E, Nkeck JR, Nyaga UF, Ngouo AT, et al. Geographic distribution of metabolic syndrome and its components in the general adult population: A meta-analysis of global data from 28 million individuals. Diabetes Res Clin Pract. 2022 Jun;188:109924.

19. Yuan S, Bruzelius M, Xiong Y, Håkansson N, Åkesson A, Larsson SC. Overall and abdominal obesity in relation to venous thromboembolism. J Thromb Haemost. 2021 Feb;19(2):460–9.

20. Sekhri T, Kanwar RS, Wilfred R, Chugh P, Chhillar M, Aggarwal R, et al. Prevalence of risk factors for coronary artery disease in an urban Indian population. BMJ Open. 2014 Dec 1;4(12):e005346.

21. Oh SK, Cho AR, Kwon YJ, Lee HS, Lee JW. Derivation and validation of a new visceral adiposity index for predicting visceral obesity and cardiometabolic risk in a Korean population. PLOS ONE. 2018 Sep 13;13(9):e0203787.

22. Kahn HS. The “lipid accumulation product” performs better than the body mass index for recognizing cardiovascular risk: a population-based comparison. BMC Cardiovascular Disorders. 2005 Sep 8;5(1):26.

23. Dobiás□ová M, Frohlich J. The plasma parameter log (TG/HDL-C) as an atherogenic index: correlation with lipoprotein particle size and esterification rate inapob-lipoprotein-depleted plasma (FERHDL). Clinical Biochemistry. 2001 Oct 1;34(7):583–8.

24. Peterson DD. History of the U.S. Navy Body Composition Program. Military Medicine. 2015 Jan 1;180(1):91–6.

25. Woolcott OO, Bergman RN. Relative fat mass (RFM) as a new estimator of whole-body fat percentage ─ A cross-sectional study in American adult individuals. Sci Rep. 2018 Jul 20;8(1):10980.

26. Krakauer NY, Krakauer JC. A New Body Shape Index Predicts Mortality Hazard Independently of Body Mass Index. PLOS ONE. 2012 Jul 18;7(7):e39504.

27. Thomas DM, Bredlau C, Bosy-Westphal A, Mueller M, Shen W, Gallagher D, et al. Relationships between body roundness with body fat and visceral adipose tissue emerging from a new geometrical model. Obesity. 2013;21(11):2264–71.

28. Valdez R. A simple model-based index of abdominal adiposity. Journal of Clinical Epidemiology. 1991 Jan 1;44(9):955–6.

29. Simental-Mendía LE, Rodríguez-Morán M, Guerrero-Romero F. The Product of Fasting Glucose and Triglycerides As Surrogate for Identifying Insulin Resistance in Apparently Healthy Subjects. Metabolic Syndrome and Related Disorders. 2008 Dec;6(4):299–304.

30. Bello-Chavolla OY, Almeda-Valdes P, Gomez-Velasco D, Viveros-Ruiz T, Cruz-Bautista I, Romo-Romo A, et al. METS-IR, a novel score to evaluate insulin sensitivity, is predictive of visceral adiposity and incident type 2 diabetes. European Journal of Endocrinology. 2018 May 1;178(5):533–44.

31. Bello-Chavolla OY, Antonio-Villa NE, Vargas-Vázquez A, Viveros-Ruiz TL, Almeda-Valdes P, Gomez-Velasco D, et al. Metabolic Score for Visceral Fat (METS-VF), a novel estimator of intra-abdominal fat content and cardio-metabolic health. Clinical Nutrition. 2020 May 1;39(5):1613–21.

32. D’Agostino RB, Vasan RS, Pencina MJ, Wolf PA, Cobain M, Massaro JM, et al. General Cardiovascular Risk Profile for Use in Primary Care. Circulation. 2008 Feb 12;117(6):743–53.

33. Selvarajah S, Kaur G, Haniff J, Cheong KC, Hiong TG, van der Graaf Y, et al. Comparison of the Framingham Risk Score, SCORE and WHO/ISH cardiovascular risk prediction models in an Asian population. International Journal of Cardiology. 2014 Sep 1;176(1):211–8.

34. Lip GYH, Nieuwlaat R, Pisters R, Lane DA, Crijns HJGM. Refining Clinical Risk Stratification for Predicting Stroke and Thromboembolism in Atrial Fibrillation Using a Novel Risk Factor-Based Approach: The Euro Heart Survey on Atrial Fibrillation. CHEST. 2010 Feb 1;137(2):263–72.

35. Tahir H, Sarwar U, Hussain A, Awan MU, Ahmad S, Ijaz MS, et al. Use of CHA2DS2-VASc Score In Patients Without Atrial Fibrillation: Review Of Literature. Journal of Cardiology & Cardiac Surgery [Internet]. 2021 Aug 3 [cited 2024 Mar 30];1(1). Available from: https://www.medijournalshub.com/article/Use-of-CHA2DS2-VASc-Score-in-Patients-without-Atrial-Fibrillation-Review-of-Literature

36. Barbar S, Noventa F, Rossetto V, Ferrari A, Brandolin B, Perlati M, et al. A risk assessment model for the identification of hospitalized medical patients at risk for venous thromboembolism: the Padua Prediction Score. Journal of Thrombosis and Haemostasis. 2010 Nov 1;8(11):2450–7.

37. Lakshmi S, Metcalf B, Joglekar C, Yajnik CS, Fall CH, Wilkin TJ. Differences in body composition and metabolic status between white UK and Asian Indian children (EarlyBird 24 and the Pune Maternal Nutrition Study). Pediatric Obesity. 2012;7(5):347– 54.

38. Lear SA, Humphries KH, Kohli S, Chockalingam A, Frohlich JJ, Birmingham CL. Visceral adipose tissue accumulation differs according to ethnic background: results of the Multicultural Community Health Assessment Trial (M-CHAT)2. The American Journal of Clinical Nutrition. 2007 Aug 1;86(2):353–9.

39. Jin J, Woo H, Jang Y, Lee WK, Kim JG, Lee IK, et al. Novel Asian-Specific Visceral Adiposity Indices Are Associated with Chronic Kidney Disease in Korean Adults. Diabetes Metab J. 2023 Mar 6;47(3):426–36.

40. Ahn SH, Lee HS, Lee JH. Triglyceride-glucose-waist circumference index predicts the incidence of cardiovascular disease in Korean populations: competing risk analysis of an 18-year prospective study. European Journal of Medical Research. 2024 Apr 2;29(1):214.

