## Supplementary Materials for "Evaluating Cardiometabolic Indices for Predicting Thrombotic Risk in the Indian Adult Population: A Comparative Analysis"

**1. FRAMINGHAM RISK SCORE**

Estimation of 10-year cardiovascular disease risk [*D’Agostino RB Sr et al.* (2008)]

**(i) Framingham Risk Score Calculation**

| **Risk Factor** | **Risk Points** | | | |
| --- | --- | --- | --- | --- |
|  | **Male** | | **Female** | |
| **Age (years)** |  | |  | |
| 30 – 34 | 0 | | 0 | |
| 35 – 39 | 2 | | 2 | |
| 40 – 44 | 5 | | 4 | |
| 45 – 49 | 7 | | 5 | |
| 50 – 54 | 8 | | 7 | |
| 55 – 59 | 10 | | 8 | |
| 60 – 64 | 11 | | 9 | |
| 65 – 69 | 12 | | 10 | |
| 70 – 74 | 14 | | 11 | |
| 75 + | 15 | | 12 | |
| **HDL-C (mmol/L)** |  | |  | |
| > 1.6 | -2 | | -2 | |
| 1.3 – 1.6 | -1 | | -1 | |
| 1.2 – 1.29 | 0 | | 0 | |
| 0.9 – 1.19 | 1 | | 1 | |
| < 0.9 | 2 | | 2 | |
| **Total Cholesterol (mmol/L)** |  | |  | |
| < 4.1 | 0 | | 0 | |
| 4.1 – 5.19 | 1 | | 1 | |
| 5.2 – 6.19 | 2 | | 3 | |
| 6.2 – 7.2 | 3 | | 4 | |
| > 7.2 | 4 | | 5 | |
| **Systolic Blood Pressure (mm Hg)** | Not Treated | Treated | Not Treated | Treated |
| < 120 | - 2 | 0 | -3 | -1 |
| 120 – 129 | 0 | 2 | 0 | 2 |
| 130 – 139 | 1 | 3 | 1 | 3 |
| 140 – 149 | 2 | 4 | 2 | 5 |
| 150 – 159 | 2 | 4 | 4 | 6 |
| 160 + | 3 | 5 | 5 | 7 |
| **Smoker** |  | |  | |
| Yes | 4 | | 3 | |
| No | 0 | | 0 | |

**(ii) 10-year Cardiovascular Disease (CVD) Risk Percentage**

| **Total Points** | **10-year CVD Risk (%)** | |
| --- | --- | --- |
|  | **Men** | **Women** |
| ≤ -3 | < 1 | < 1 |
| -2 | 1.1 | < 1 |
| -1 | 1.4 | 1.0 |
| 0 | 1.6 | 1.2 |
| 1 | 1.9 | 1.5 |
| 2 | 2.3 | 1.7 |
| 3 | 2.8 | 2.0 |
| 4 | 3.3 | 2.4 |
| 5 | 3.9 | 2.8 |
| 6 | 4.7 | 3.3 |
| 7 | 5.6 | 3.9 |
| 8 | 6.7 | 4.5 |
| 9 | 7.9 | 5.3 |
| 10 | 9.4 | 6.3 |
| 11 | 11.2 | 7.3 |
| 12 | 13.3 | 8.6 |
| 13 | 15.6 | 10.0 |
| 14 | 18.4 | 11.7 |
| 15 | 21.6 | 13.7 |
| 16 | 25.3 | 15.9 |
| 17 | 29.4 | 18.51 |
| 18 | >30 | 21.5 |
| 19 | >30 | 24.8 |
| 20 | >30 | 27.5 |
| 21+ | >30 | >30 |

**(iii) CVD Risk Stratification**

| **Risk Level** | **10-year CVD Risk %** |
| --- | --- |
| Low | < 10 |
| Intermediate | 10 – 19 |
| High | ≥ 20 |

**Note:** Coronary heart disease, diabetes mellitus, carotid artery disease, peripheral arterial disease, abdominal aortic aneurysm, and chronic kidney disease are considered coronary heart disease risk equivalents with a 10-year CVD risk of 20% (NCEP ATP III guidelines).

**2. CHA_2_DS_2_-VASc SCORE**

Assessment of risk of stroke in patients with non-valvular atrial fibrillation [*Lip et al.* (2010)]

**(i) Risk Score Calculation**

| **Risk Factor** | **Score** | **Definition** |
| --- | --- | --- |
| **C**ongestive Heart Failure/LV dysfunction | 1 | Left ventricular dysfunction or symptomatic heart failure |
| **H**ypertension | 1 | More than 140/90 mmHg (use of 130/80 mmHg is acceptable) or on anti-hypertensive therapy |
| **A**ge ≥ 75 years | 2 |  |
| **D**iabetes Mellitus | 1 | Fasting blood glucose > 126 mg/dL, HgA1c > 6.5%, or receiving treatment for diabetes |
| **S**troke/TIA/TE | 2 | Prior history of stroke, TIA, or systemic embolism |
| **V**ascular Disease | 1 | Prior myocardial infarction (MI), angina pectoris, percutaneous coronary intervention or coronary artery bypass surgery, intermittent claudication, previous surgery or percutaneous intervention of the abdominal aorta or lower extremity vessels, abdominal or thoracic surgery, arterial and venous thrombosis |
| **A**ge 65 – 74 years | 1 |  |
| **S**ex **C**ategory (Female) | 1 |  |

**(ii) Risk Stratification**

| **Risk Level** | **Total Score** |
| --- | --- |
| Low-risk | 0 |
| Medium-risk | 1 |
| High-risk | ≥ 2 |

**3. PADUA PREDICTION SCORE**

Evaluation of risk of venous thromboembolism in non-surgical hospitalized patients and determination of the need for pharmacological prophylaxis [*Barbar et al.* (2010)].

**(i) Risk Score Calculation**

| **Baseline Features** | **Score** |
| --- | --- |
| Active cancer (Patients with local or distant metastases and/or in whom chemotherapy or radiotherapy had been performed in the previous 6 months) | 3 |
| Previous VTE (with the exclusion of superficial vein thrombosis) | 3 |
| Reduced mobility [Bedrest with bathroom privileges (either due to patient’s limitations or on physician’s order) for at least 3 days] | 3 |
| Already known thrombophilic condition (Carriage of defects of antithrombin, protein C or S, factor V Leiden, G20210A prothrombin mutation, antiphospholipid syndrome) | 3 |
| Recent (<1 month) trauma and/or surgery | 2 |
| Elderly age (≥ 70 years) | 1 |
| Heart and/or respiratory failure | 1 |
| Acute myocardial infarction or ischemic stroke | 1 |
| Acute infection and/or rheumatologic disorder | 1 |
| Obesity (BMI ≥ 30) | 1 |
| Ongoing hormonal treatment | 1 |

**(ii) Risk Stratification**

| **Risk Level** | **Total Score** |
| --- | --- |
| Low-risk | < 4 |
| High-risk | ≥ 4 |
